# Perceptions of gender equity and markers of achievement in a National Institute for Health Research (NIHR) Biomedical Research Centre (BRC): A qualitative study

**DOI:** 10.1101/2021.07.29.21261318

**Authors:** Lorna R Henderson, Rinita Dam, Syed Ghulam Sarwar Shah, Pavel V Ovseiko, Vasiliki Kiparoglou

## Abstract

**Background:** The need to improve gender equity (GE) in academic medicine is well documented. Biomedical Research Centres (BRCs), partnerships between leading National Health Service (NHS) organisations and universities in England, conduct world-class translational research funded by the National Institute for Health Research (NIHR). In 2011, eligibility for BRC funding was restricted to universities demonstrating sustained GE success recognised by the Athena SWAN Charter for Women in Science Silver awards. Despite this structural change, GE research in BRC settings is underdeveloped, yet critical to the acceleration of women’s advancement and leadership.

**Objectives:** To explore both women’s and men’s perceptions of GE and current markers of achievement in a BRC setting.

**Methods:** Thematic analysis of data from two discrete research projects: 53 GE survey respondents’ free text comments (34 women, 16 men), and 16 semi structured interviews with women affiliated to the NIHR Oxford BRC.

**Results:** Four major themes emerged from the analysis: perceptions of the Athena Swan Charter for Women in Science (GE policy); views on monitoring GE in BRCs; views on current markers of achievement in academia and GE; and recommendations for actions to improve GE in BRC settings. Monitoring of GE in BRCs was deemed to be important, but complex. Participants felt current markers of achievement were not equitable to women as they did not take contextual factors into account such as maternity leave and caring responsibilities. BRC specific organisational policies and metrics are required to monitor and catalyse GE.

**Conclusions:** Markers of achievement for monitoring GE in BRCs should take into account contextual factors specific to BRCs and women’s career progression and professional advancement. GE markers of achievement should be complimented with broader aspects of equality, diversity and inclusion.

## Introduction

Women are underrepresented in senior leadership positions in academic medicine settings compared to men (1–8). Yet, gender equity (GE) is widely recognised to be an important driver of successful competitive organisations (4, 8–10) Furthermore, the European Commission recently set targets to increase the representation of women on decision making bodies to at least 30%-40% referred to as the “gender balance zone” (10, 11). In this study, when referring to GE we adopt the UNESCO definition: “fairness of treatment for women and men, according to their respective needs. This may include equal treatment or treatment that is different which is considered equivalent in terms of rights, benefits, obligations and opportunities” (12).

In England, demonstrating sustained GE improvements in BRCs has been a required indicator to apply for National Institute for Health Research (NIHR) translational research funding (4, 6,13–14). In 2011, the Chief Medical Officer, Dame Sally Davies, announced only medical schools holding the Silver Award of the Athena Swan Charter for Women in Science award (denoting significant achievements, impact and evidence in GE) would be eligible to apply for BRC funding (6, 13). However, whilst Athena SWAN awards have been important catalysts for GE in University settings, they were not designed for translational research organisations (TROs) such as BRCs (4,6,14). There is a recognised research gap regarding GE in BRC settings as studies are typically set in Universities (4–6). This study sets out to address this gap.

In 2020, the NIHR ended the specific requirement for BRCs to hold Athena Swan Charter Silver awards to be eligible for funding (15). Their recent Equality, Diversity and Inclusion (EDI) strategic plan now requires BRCs to provide evidence of such activities at all levels, not only GE, to be eligible to apply for translational research funding (15).

### Study Objectives

This study sets out to explore both women’s and men’s perceptions of the importance of monitoring and measuring GE and current markers of achievement in an NIHR BRC. The aim is to create context specific evidence for NIHR BRCs to facilitate women’s advancement and leadership progression in translational research (4, 6). Further details can be found in the study protocol (4).

### Biomedical Research Centres

BRCs are partnerships between the UK’s leading NHS organisations and universities. There are currently 20 BRCs in England, that together have been awarded significant funding (£816 million commencing 2017 to 2022) to conduct translational research by world class researchers to develop innovative treatments for patient benefit (15–16).

### Study setting

This study was conducted at the NIHR Oxford BRC - a leading translational research organisation based at the Oxford University Hospitals NHS Foundation Trust and run in partnership with the University of Oxford (17). It is one of 20 NIHR BRCs in England and in 2016 was awarded £113.7m for the period from 2017 to 2022 to support translational research. The NIHR Oxford BRC is divided into 20 research themes comprising four clusters: Precision Medicine, Technology and Big Data, Immunity and Infection, and Chronic Diseases (17, 18).

## Methods

The study is based on the thematic analysis of qualitative data from two discrete research projects: (1). 53 free-text responses (34 women, 16 men, 2 preferred not to say, 1 self-described), from a larger GE survey conducted at the NIHR Oxford BRC (2, 6). 16 separate semi-structured interviews were conducted with a sample of women affiliated to the NIHR Oxford BRC.

### Free text responses

The free text comments formed part of an online GE survey distributed to senior leadership, clinical and non-clinical researchers, trainees, administrative and other professionals affiliated to the NIHR Oxford BRC (N = 683) from May to July 2019 (6). In addition to quantitative questions, the analysis of which is reported elsewhere (6), the survey contained two open ended questions, which are analysed in this paper. Participants were asked to share their views on (1) “other indicators related to gender equity that the BRC should assess and monitor” and (2) “comments or suggestions on new ways of measuring gender equity in Biomedical Research Centres”.

### Semi-structured interviews

The semi-structured interviews were completed from August 2018 to February 2019. The interviewees included a purposive sample of women affiliated to the NIHR Oxford BRC at different careers stages (early-career researchers, postdoctoral researchers, and professors) across a range of departments and disciplines. Senior female leaders and managers were also invited to encourage a diverse range of respondents. An email invitation was sent to all BRC research theme managers with an information sheet concerning the study and a brief overview of the aims of the study. A snowballing recruitment strategy was also adopted (18–19)

Recruitment of interviewees continued until no significant new themes were emerging from the interviews (19–20). Semi-structured interviews were conducted by the lead author (LH), a trained qualitative researcher with a background in medical sociology. Participants were provided with an information sheet and asked to sign a consent form.

A brief introduction on the scope of the interview and research project was provided at the beginning of the interview. Interviews were tape recorded and transcribed verbatim.

Transcripts were returned to the interviewees to approve. The length of the interviews ranged from thirty to sixty minutes. The majority of interviews took place in the interviewee’s workplace in a confidential setting. The remainder were conducted in the researcher’s office at the participants’ request. The interview schedule was informed by a literature review and was inductive.

The key areas covered were views on:

- The Athena Swan Charter for Women in Science
- Monitoring GE in BRCs
- Current markers of achievement and GE
- New ways of capturing GE in BRCs
- Recommendations for actions to improve GE in BRC settings

Participants were encouraged to make additional comments and questions were adapted to ensure relevance for respective participants.

### Ethics statement

The study was reviewed by the Officer of the Oxford University Medical Sciences Inter-divisional Research Ethics Committee and the University of Oxford Clinical Trials and Research Governance Team who determined that the study was exempt from full ethical review.

### Data analysis

The free text survey comments were analysed by LH, SGSS and RD. Thematic analysis was carried out by the qualitative research team (LH and RD) (21). To establish trustworthiness (22), LH and RD independently read the transcripts line by line, identifying emergent themes and created initial codes. LH and RD brought codes together to create a coding framework and coded the transcripts with NVIVO 11 (23). LH and RD conducted constant comparison, an iterative method of analysis, searching for each themed code throughout the entire data set and comparing all instances until no new themes were identified. Emerging findings were discussed at team meetings to resolve discrepancies and refine themes. Divergent views and areas of diversity were considered. Researchers used relevant studies against the analysis to check emerging themes (19, 20). Prior experiences and views, which may have influenced the analysis were discussed. The final stage involved selecting quotes to illustrate major themes and the diversity of responses.

## Findings

53 (22%) of 243 GE survey respondents, provided free text comments (34 women, 16 men, 2 preferred not to say, 1 self-described), and 16 separate semi structured interviews with a purposive sample of women affiliated to the NIHR Oxford BRC. Demographic characteristics of the survey participants and interviewees are presented in Table 1 and 2. The findings are based on the combined thematic analysis of 53 free-text responses of GE survey respondents and semi-structured qualitative interviews (n=16). Data analysis identified four main themes and twelve corresponding sub-themes. Table 3 presents an overview of the coding structure. The main themes were: 1. views on the Athena Swan Charter for Women in Science, 2. views on monitoring GE in BRCs, 3. views on current markers of achievement in BRCs and GE, 4. recommendations for actions to improve GE in BRC settings. The themes are presented in detail in the next section together with illustrative quotes. The findings are reported in line with the COREQ guidance (24). Illustrative quotes are presented with relevant demographic data for gender (female=F, male=M, prefer not to say=X), BRC affiliate category and the data source (GE Survey=GES or qualitative interviews= QI) (see Table 1 and 2).

**Table 1.** Demographic characteristics of participants from GE Survey.

| Characteristics | N= 53 |
| --- | --- |
| **Sex** | n (%) |
| Female | 34 (64%) |
| Male | 16 (30%) |
| Prefer to self describe | 1 (2%) |
| Prefer not to say | 2 (4%) |
| **Age** |  |
| 18-30 years | 3 (6%) |
| 31-40 years | 13 (25%) |
| 41-50 years | 19 (36%) |
| 51-60 years | 13 (25%) |
| 61 + years | 3 (6%) |
| Prefer not to say | 2 (4%) |
| **BRC Affiliate Category** |  |
| **Investigator** e.g. Principle Investigator/Co-Investigator/ Chief Investigator | 20 (38%) |
| Research Associate (e.g. Researcher and Research Fellow) | 12 (23%) |
| Admin/Technical/Professional manager/Support | 15 (28%) |
| Other | 4 (4%) |
| Prefer not to say | 2 (4%) |
| **Duration of Affiliation to the BRC** |  |
| Up to 2 years | 15 |
| 3-7 years | 21 |
| More than 7 years | 14 |
| Prefer not to say | 2 |
| Missing information | 1 |

**Table 2.** Demographic characteristics of participants from GE Qualitative Interviews.

| Characteristics | GE Qualitative Interviews (N=16)<br>n (%) |
| --- | --- |
| **Sex** |  |
| Female | 16 (100) |
| **BRC Affiliate Category** |  |
| Early career researchers | 3 (18.8) |
| Senior postdoctoral researchers | 4 (25) |
| Associate Professors | 4 (25) |
| Professors | 1 (6.3) |
| Senior Managers | 3 (18.8) |
| Manager | 1 (6.3) |

**Table 3.** Description of the coding tree

| Main Themes | Sub themes |
| --- | --- |
| **1. Views on the Athena SWAN Charter for Women in Science** | • Catalyst for change<br>• Limitations of Athena SWAN<br>• Additional organisational support for those with childcare responsibilities required |
| **2. Views on monitoring GE in Biomedical Research Centres** | • Important to monitor GE in BRC settings<br>• Complexity of monitoring GE<br>• Broader equality, diversity and inclusion |
| **3. Views on current markers of achievement and GE** | • Context is important<br>• Perceptions of structural barriers to GE<br>• Concerns about positive discrimination |
| **4. Recommendations for actions to improve GE in BRC settings** | • Monitor BRC GE metrics at an organisational level<br>• Monitor BRC recruitment and retention by gender<br>• Monitor academic citizenship activities by gender<br>• Create BRC GE organisational processes to catalyse sustainable change in GE |

### Views on the Athena SWAN Charter for Women in Science

#### Catalyst for change

Several interviewees described how the Athena Swan GE Charter for Women in Science link to NIHR BRC funding eligibility had catalysed positive change in GE. For example, increasing the diversity of committee membership, and changing the timing of department meetings to take participants’ caring responsibilities into account. One senior academic described the benefits of changing meeting times as a consequence of Athena SWAN:

> *“I do know about Athena SWAN…People do moan about it but I think it genuinely has made a difference…Very, very simple things actually make a big difference, like moving meetings to times…in the middle of the day so you can go and don’t have all the discussions that are interesting in the pub afterwards because I can’t do that…”* (QI, 11, F, Associate Professor)

#### Limitations of Athena SWAN

Conversely, several interviewees described the Athena SWAN Charter as a “box ticking exercise” implemented primarily because of the link to NIHR BRC research funding and questioned whether it had led to sustainable change in women’s research careers. Athena SWAN committees were described as overtly time consuming and bureaucratic with discussions predominantly focussed on women’s childcare responsibilities not career progression. Strong commitment by senior leadership was required to catalyse sustainable change in GE as this senior researcher explained:

> *“Athena SWAN exists is so that we’re eligible for things like NIHR funding. If you really were interested in equality, then you would go to the very top of people in divisions and make them deal with gender bias, not coming and putting more workload on people like me who are already affected by it and have precious enough little time to do things like write grants as it is”*. (QI,14, F Associate Professor).

#### Additional organisational support for those with childcare responsibilities required

Survey respondents identified the need for additional organisational support for those with childcare responsibilities. Participants wanted dedicated funds to support maternity leave, childcare costs and caring responsibilities. Several respondents described how timings of departmental events and meetings did not always take those with childcare responsibilities into account:

> *“Participation in departmental seminars / workshops: often these are timed to go on beyond the end of the working day, which excludes anyone with childcare responsibilities (predominantly women/early career researchers) from fully participating*.*”* (GES R228, F, Admin Staff Member)

### Views on monitoring GE in BRCs

#### Important to monitor GE in BRC Settings

Survey and interview participants commented it was highly important to monitor and benchmark GE in BRCs. This was felt to be particularly important in clinical academic medicine where representation of women is traditionally low as this senior female investigator described:

> *“The gender imbalance is particularly noticeable in clinical rather than non-clinical staff and this must be monitored. At present there is very little information on this and therefore ways to address the issues*.*”* (GES R243, F, Principal Investigator)

#### Complexity of monitoring GE

Whilst participants highlighted monitoring gender equity was extremely important through benchmarking of data. Others highlighted the complexity of gathering such data. This industry manager highlighted gender and industry metrics are not routinely monitored and assigning gender to data would be challenging:

> *“I have never recorded gender against anything, apart from putting someone’s name…Nowadays you wouldn’t want to assume somebody’s gender either so you couldn’t judge it wholly on someone’s name… It’s not something we record so it would be very difficult to report on it I think*.*”* (QI 2, F, Manager).

#### Broader equality, diversity and inclusion

Both male and female interviewees and survey respondents felt it was important to monitor not only GE (which is the remit of the Athena SWAN Charter), but also characteristics of diversity:

> “*Other aspects of diversity are as important as gender and also need to be monitored. Specifically disability, original social class, (and) ethnicity. The clinical research community (in and out of Oxford) is remarkably non diverse when this broader aspect beyond Athena SWAN is considered and is not representative of the NHS workforce diversity*.*”* (GES R96 M, Other)

Survey respondents suggested monitoring GE while taking into account the intersectionality of gender with other aspects of identity:

> “*Intersections between gender and other factors associated with oppression, such as race, sexuality and transgender identities*.*”* (GES R140, Prefer to Self describe, Support Associate).

### Views on current markers of achievement and GE

#### Context is important

Many survey and interview participants described the limitations of current markers of achievement because they lacked important contextual adjustments e.g. career breaks for maternity care, part time working, and caring responsibilities. As this female senior manager highlighted, absolute numbers for certain markers of achievement such as peer reviewed publications were not necessarily equitable to women who had taken maternity leave:

> *“Context is important… obviously the number of publications – that’s relatively easy…that can be a little bit nuanced as well because that may be in the context of having maternity leave one year. So parental leave is quite important - this applies to men too. I don’t think this is necessarily completely focussed on women because you need that comparator group…Maternity leave, output, grant applications, whether they are full time or part time, and also I think qualitative data here is quite important too because I don’t think that statistics on their own can tell the whole story*.*”* (QI 4, F, Senior Manager).

#### Perceptions of structural barriers to GE

Others felt disadvantaged when applying for research grants and promotion which did not take into account maternity leave when assessing their academic track record:

> *“ I feel I was always a bit more delayed in the career progression than male counterparts. Many times grant bodies didn’t take that into account…if you have had maternity leave…The guidelines to be a university research lecturer… are the same for male and female but you cannot really measure the experience of a female researcher the same way as you measure a male researcher. It is quite different. You only have to look around, many of the PIs (Principal Investigators) are males and the postdocs are females. Why is that*?” (QI6, F, Investigator)

#### Concerns about positive discrimination

Conversely, several survey respondents and interviewees raised concerns about positive discrimination stating they did not wish to be promoted simply because of their gender but rather due to their contribution to science as this senior researcher described:

> *“I didn’t want to get anything because I was female. I wanted to get it because I deserved it…The whole point is this person can do the job whoever they are and they might do the job differently because they are female…it’s about being capable within that role*.*”* (QI 7, F, Associate Professor)

### Recommendations for actions to improve GE in BRC settings

#### Monitor GE metrics at an organisational level

Both survey and interview respondents raised the importance of monitoring metrics by gender at an organisational level in a range of specific areas, and they proposed a range of actions from benchmarking the career development of female early career researchers and overall career progression within the BRC.

Similarly, early career researchers felt it was important to monitor success in research by gender at a department level so that any inequities could be better understood. This senior female investigator flagged up the importance of monitoring gender and awards:

> *“For any BRC post or call for funding scheme, please publish how many male/female applicants were received, and how many…achieved the post/award…Merit/track record should be the primary assessment criterion. It would also be concerning if only one sex/gender tends to predominate the winners list*.*”* (GES R273, F, Investigator)

#### Monitor BRC recruitment and retention by gender

Participants felt it was also important to monitor recruitment and retention for GE and proposed a range of actions. These included monitoring the seniority of staff and gender and their job roles to assist exploring retention and recruitment processes, gender balance of interview panels and benchmarking number of applicants for posts by gender:

> *“Compare number of senior investigators and Professors with the numbers and sex of postdocs and doctoral students, where are people dropping out? or where are the recruitment practices potentially biased?”* (GES R227, M, Research Associate)

#### Monitor academic citizenship activities by gender

Female interviewees described the importance of recognising teaching, peer review and committee work often referred to as “the housework of academia”. These highly administrative and important but time consuming activities often took time away from their core academic activities, e.g. writing grants or peer reviewed publications but were not recognised as “markers of achievement”. Ironically, due to underrepresentation of women in some departments some women felt more obliged to fulfil these roles compared to their male colleagues as this senior female academic describes:

> *“The housework of academia…is under-recognised and I think…women end up doing more of it. If it didn’t happen, the university would not continue to run…Committee memberships is difficult and teaching… It’s the softer stuff…People realising that they need to have women helping out on committees and doing this is brilliant but if there aren’t actually that many women, you do get asked to do it a lot*… *Whereas I think, for whatever reason, men are better at working out what’s going to get them where they want to be and do that”*. (QI, 11, F, Associate Professor)

#### Create BRC GE organisational policies to catalyse change

Male and female survey respondents and interviewees raised the importance of specific organisational policies at an institutional level to support career progression irrespective of gender. Some respondents stated that gender diversity in senior leadership roles would demonstrably improve GE. Inequity in pay was also raised by participants as an important marker of GE. Organisational processes and policies should be implemented to support GE as this female researcher describes:

> *“You should create processes that remove bias and honour achievement, irrespective of gender or other identifiers”*. (GES R225, F, Research Associate)

## Discussion

### Key findings

Participants perceived linking the Athena SWAN Charter for Women in Science to eligibility for BRC funding to have been an important driver for GE for BRC affiliates with positive changes in working practices at departmental level. However, despite the implementation of Athena SWAN, many participants still flagged up the need for additional support for those with childcare responsibilities including dedicated funds to cover such costs.

Furthermore, some participants described it as a bureaucratic “box ticking” exercise dominated by all female committees. This is consistent with previous research highlighting Athena SWAN Charter processes to be highly administrative, and where committees are dominated by women this may inadvertently reproduce gender inequity rather than address it (5). Furthermore improving GE takes significant time and is neither academics primary role nor typically rewarded (25). Previous research has analysed the Athena SWAN Charter in light of complexity, including the structural, institutional and cultural biases towards women’s careers arguing accordingly complex interventions are required to drive GE improvements (26).

The majority of both male and female respondents felt monitoring and measuring GE in BRC settings was very important but complex. Participants felt current markers of achievement were not equitable to women as they did not take contextual factors into account e.g. maternity leave, part time working and carers leave. Participants recommended a range of new areas to monitor GE and organisational policies specific to BRCs to support GE including senior leadership and institutional support. This finding is consistent with a recent survey of markers of achievement in a BRC where participants ranked BRC senior leadership roles and organisational policies on GE to be the most important indicators (6). BRC specific measures are necessary because BRCs are co-hosted by a University and an NHS Trust with separate practices and policies on GE. Despite the implications of Athena SWAN Awards for BRC funding, assessment for awards has focused only on practice in University settings (5, 6). Female participants described the burden of academic “citizenship” work, often referred to in the literature as “the housework of academia” e.g. time consuming committee work, teaching and highly administrative work not recognised as a marker of achievement yet central to academic life. This finding is consistent with previous research which has described such work as a burden and barrier to progression for women in universities arguing it should be equally valued as research in order to address pay inequalities (27–31). Being able to negotiate academic housework is important, with the amount taken on being linked to power structures in academia (30). As in the current study, previous research has identified where women are underrepresented in organisations, they felt more obliged to fulfil these roles e.g. committee membership and mentorship compared to male colleagues (30, 31).

Participants suggested monitoring broader aspects of EDI, especially disability, social class, ethnicity, career stages as well as broader indicators such as race, sexuality and transgender identities. This is particularly relevant to the given biomedical research setting given the NIHR commitment to all aspects of EDI (15). This can also help to address GE more broadly, taking into account intersectional considerations of other forms of identity intersectionaly. This is also timely given the recognition of the importance of diversity and inclusivity for biomedical research organisations more broadly (35). In the United States, the National Institutes of Health (NIH) have also committed to increase the diversity of the biomedical research workforce (36).

### Recommendations

Current academic markers of achievement in BRCs reward absolute numbers of research outputs e.g. peer reviewed publications, citations, grants and intellectual property (5). Despite the drive to improve GE in BRCs, these are not adjusted for women taking time out for maternity leave or part time working. Furthermore data on GE is not routinely monitored or benchmarked in BRCs, and it is complex to collate (4, 6). As identified in the literature multiple complex factors contribute to the slow pace of women’s advancement into leadership positions in academic medicine (4, 24). GE initiatives may be impacted by Institutional, national and societal issues (6). Participants felt monitoring GE was important but contextual factors must be taken into account when comparing research outputs. Taking the academic citizenship role into account in terms of promotion is key (23). Whilst Athena SWAN is an important driver for GE, more localised organisational policies specific to the BRC and staff who are employed by the NHS are required. This particularly concerns monitoring broader aspects of EDI, especially disability, social class and ethnicity and broader indicators such as race, sexuality and transgender identities.

### Strengths and limitations

The main strength of this research is this is the first qualitative study to our knowledge exploring women’s and men’s perceptions of GE specifically in an NIHR Oxford BRC setting. Another strength is that we have combined two different data sets. Using two different data sets broadens the diversity of responses to views on GE including men’s perspectives. This approach has been successfully applied in previous exploratory research on Athena SWAN (32).

Notably, the responses to the survey were anonymous and so this may have enabled more critical views to be provided. Limitations of the research are the qualitative interview sample is relatively small and only includes women; however, it is exploratory and the results can be used to inform future research in this field. Typically qualitative interviews provide richer data which is key when exploring under-researched topics (19) such as GE in NIHR BRC settings.

It is therefore acknowledged that the results may not be fully generalisable to other BRCs and the entire population given that BRCs cover both hospital and university settings with a broad range of clinical, non-clinical and research staff at all levels. Future research is needed to determine whether the results hold true in other BRCs. A further limitation is we did not collect information on ethnicity and other aspects of EDI. This is an important dimension to include in future work to extend the focus of GE to address understanding of EDI within a BRC given the prioritisation of this work by their funder NIHR (15).

## Conclusions

Gender inequity in academic medicine is well documented as an area to be addressed. This is one of the first studies to explore women’s and men’s views on markers of achievement for women in academic science specifically in a BRC. Previous research in this field has focussed predominantly on Athena SWAN initiatives in universities whereas this paper has a broader remit (5). This study contributes towards BRCs’ need to extend understanding of gender equity to facilitate the acceleration of women’s advancement and leadership in translational research (5, 6). The study also draws attention to monitoring broader aspects of EDI in biomedical research settings. Given the significant investment in BRCs this is of particular relevance to the translational research workforce, patients and the public (15).

## Data Availability

Copy of questionnaire available on request

## Contributors

LH is the corresponding author, conducted the interviews and drafted the manuscript. LH, RD and SGSS contributed to the analysis and interpretation of the data. All of the authors read, checked and approved the final version of the manuscript.

## Ethics approval

The study was reviewed by the Officer of the Oxford University Medical Sciences Inter-divisional Research Ethics Committee and the University of Oxford Clinical Trials and Research Governance Team who determined that the study did not require full ethical review.

## Disclaimer

The views expressed are those of the author(s) and not necessarily those of the NHS, the NIHR or the Department of Health.

